# Exploring Multilevel Determinants of Stillbirth: A Comprehensive Analysis Across Sub-Saharan African Countries

**DOI:** 10.1101/2024.05.16.24307507

**Authors:** Khadijat Adeleye, Oluwabunmi Ogungbe, Yvette Yeboah-Kordieh, Ashley Gresh, Favorite Iradukunda

## Abstract

**Background:** Stillbirths and associated outcomes remain a significant concern in Sub-Saharan Africa (SSA), with approximately 44% of global stillbirths.

**Methods:** Using Demographic and Health Surveys (DHS) data, this study examined determinants of stillbirth among women in 29 SSA countries. Our cross-sectional analysis included a weighted sample of women 15-49 years of age who had given birth or experienced stillbirth. We used bivariate analyses and multilevel logistic regression approaches.

**Findings:** Stillbirth prevalence was 319·06/1000 live births. Among individual-level factors, risk increased with age. Higher maternal education levels were significantly associated with decreased stillbirth risk. Single women had significantly lower odds of stillbirth compared to those who no longer lived together/separated from their partner. Contextually, women with a job had an increased risk compared to women without a job, and living in a rural residential area was a significant factor.

**Interpretation:** The complex interplay of individual-level factors and contextual factors influences stillbirth outcomes in SSA. Cross-sector holistic approaches to maternal and neonatal health are needed to address the multifaceted determinants of stillbirths.

**Funding:** There was no funding for the study.

**Evidence before the study:** The prevalence of stillbirth is higher in SSA compared to other regions. Factors influencing stillbirth are complex and include individual, household, and community-level factors. We searched PubMed and Medline with no language restrictions using the search terms (“stillbirth” AND “determinant” AND “Sub-Saharan Africa”). By 2023, no studies were published on the determinants of stillbirth in Sub-Saharan Africa. Earlier studies were conducted in the context of other countries without using the calendar method to calculate stillbirth or the national demographic dataset.

**Added value of this study:**

- The relative importance of risk factors for stillbirth in different SSA countries.
- Protective effect of household leadership dynamics on reducing stillbirth odds in SSA.

**Implications of all the available evidence:**

- Focused interventions to reduce stillbirths, such as promoting female household leadership and equity.
- Improving access to education and maternal health literacy.
- Public health initiatives to prioritize social and familial support for pregnant women to create environments conducive to positive pregnancy outcomes.
- Clinicians could promote pregnancy spacing and family planning to promote optimal maternal and child health, especially among women with higher parity.
- Healthcare policies for more investment and strengthening of maternal and child care services.

## Background

Despite considerable progress in reducing maternal and neonatal mortality worldwide, stillbirth rates remain alarmingly high, particularly in low- and middle-income countries (LMICs), with sub-Saharan Africa (SSA) bearing a disproportionate burden. Stillbirth is a significant public health concern with profound emotional, social, and economic implications for affected individuals and communities. It encompasses fetal death from pregnancies starting at 28 weeks. ^1^ In SSA, perinatal mortality rates (stillbirths and early neonatal deaths) range from 34·7 to 58·4 per 1000 live births, accounting for 41% of total newborn deaths globally.^2,3^ The persistence of high stillbirth rates in SSA underscores the need to understand the multifaceted determinants contributing to this devastating outcome.

Factors driving stillbirth can be considered through a socioecological lens, which analyzes linkages among individual, household, community, and societal influences on health outcomes.^2,4,5^ Individual factors are shaped by sociocultural norms, health literacy, and access to healthcare services, emphasizing the impact of micro- and meso-system interactions on stillbirth rates.^6–8^ At the household level, the framework considers interactions between family members and their immediate environment. ^5,7^

At the community level, the broader sociocultural context and access to healthcare services impact stillbirth rates. Community norms, beliefs, and practices around childbirth and pregnancy care can shape maternal behaviors and influence prenatal care utilization.^9^ Healthcare infrastructure, availability of skilled birth attendants, and distance to healthcare facilities are crucial in determining access to timely and quality care.^10^ Community factors cause disparities in access to maternal health care services, including antenatal, intrapartum, and postnatal care. ^11,12^ These disparities increase the risks of complications and stillbirth.

Current research on stillbirth determinants in SSA countries focuses on individual factors like maternal age, education, and parity. This research is often limited to a few countries (Ethiopia, Kenya, Nigeria).^13,14^ There is little evidence regarding household and community-level contributions to stillbirth in other SSA countries. To address this gap, we used the Demographic and Health Surveys (DHS) to investigate the determinants of stillbirth among women in SSA who have been pregnant. Our study examined the multilevel factors contributing to the risk of stillbirth. Hypothesis: Individual-level risk factors for stillbirth, such as maternal age, parity, and underlying health conditions, interact with contextual factors, such as access to healthcare services, socioeconomic status, and household dynamics

## Methods

### Study design and overview

Hypothesis: Individual-level risk factors for stillbirth, such as maternal age and parity, interact with contextual factors, such as access to healthcare services, socioeconomic status, and household dynamics.

The study drew from a retrospective dataset from the DHS, collected every five years using pretested validated quantitative tools and structured methodologies.^15^ The DHS’s two-stage sampling procedure consists of a primary survey unit; participants are randomly selected from clusters in each country.^16^ The DHS sampling methodology has been published in the literature.^17^ The survey method’s consistency over time and across countries made a multi-country analysis possible. DHS datasets used in this study are publicly available on the DHS website and can be downloaded for free upon request via https://dhsprogram.com/data/available-datasets.cfm.

Countries lacking the variable (v-cal) for stillbirth calculation, including Cameroon, Congo, Cote d’Ivoire, Chad, and Togo, were excluded from the analysis (**Figure 1**). We extracted this study’s data from the most recent nationally representative DHS iterations of 29 SSA countries with information on SSA birth histories between 2010 and 2019 (**Figure 2**). The sample represents 59% of SSA countries (29/49). We pooled the data from individual recode files in each country that reported at least one birth in the five years before the surveys and included a live-term birth or stillbirth. Our total sample was 197,329; samples for specific countries are outlined in **Table 1**. Our reporting is guided by the Strengthening Reporting of Observational Studies in Epidemiology (STROBE) guidelines.^18^

**Figure 1.**
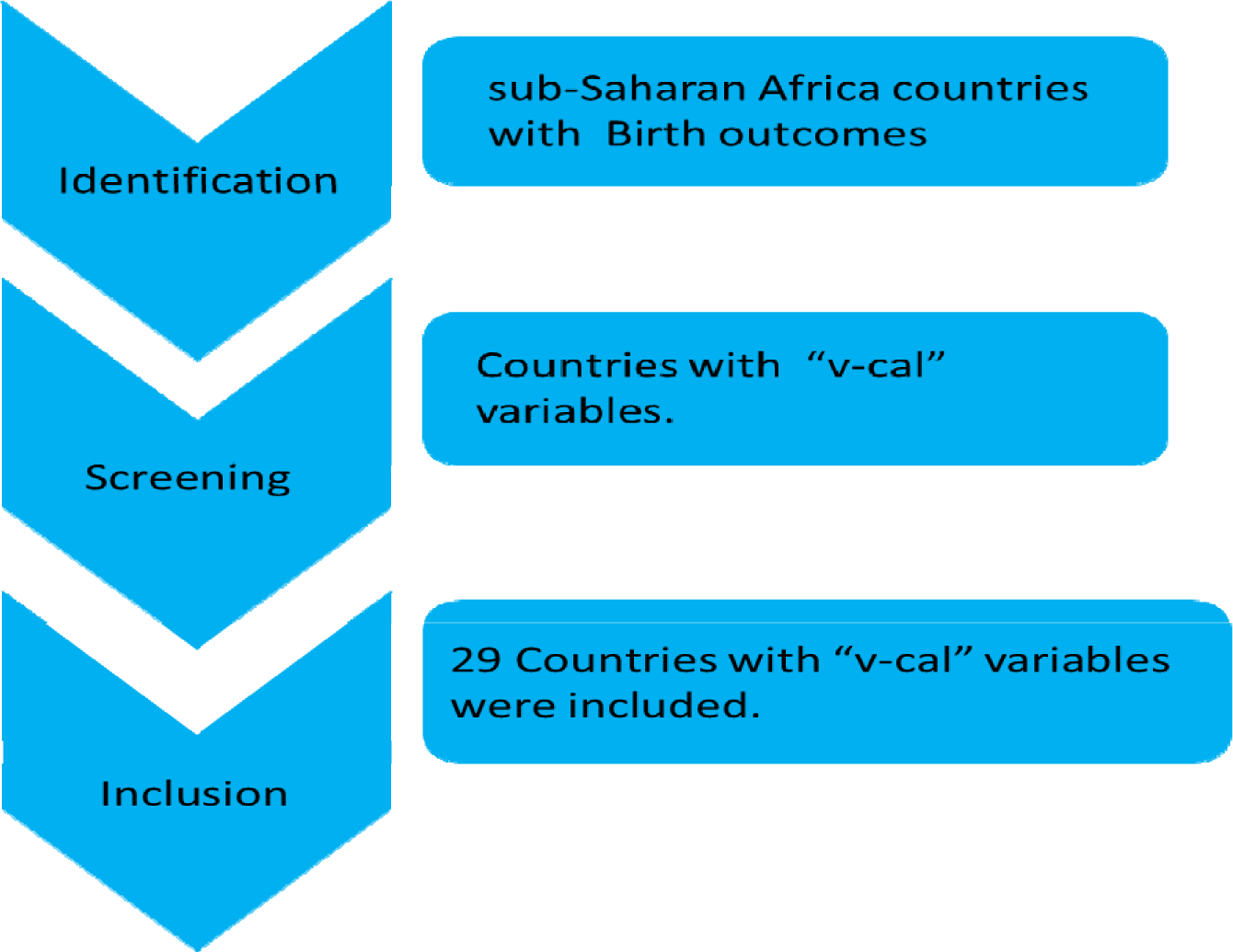
Study eligibility criteria.

**Figure 2.**
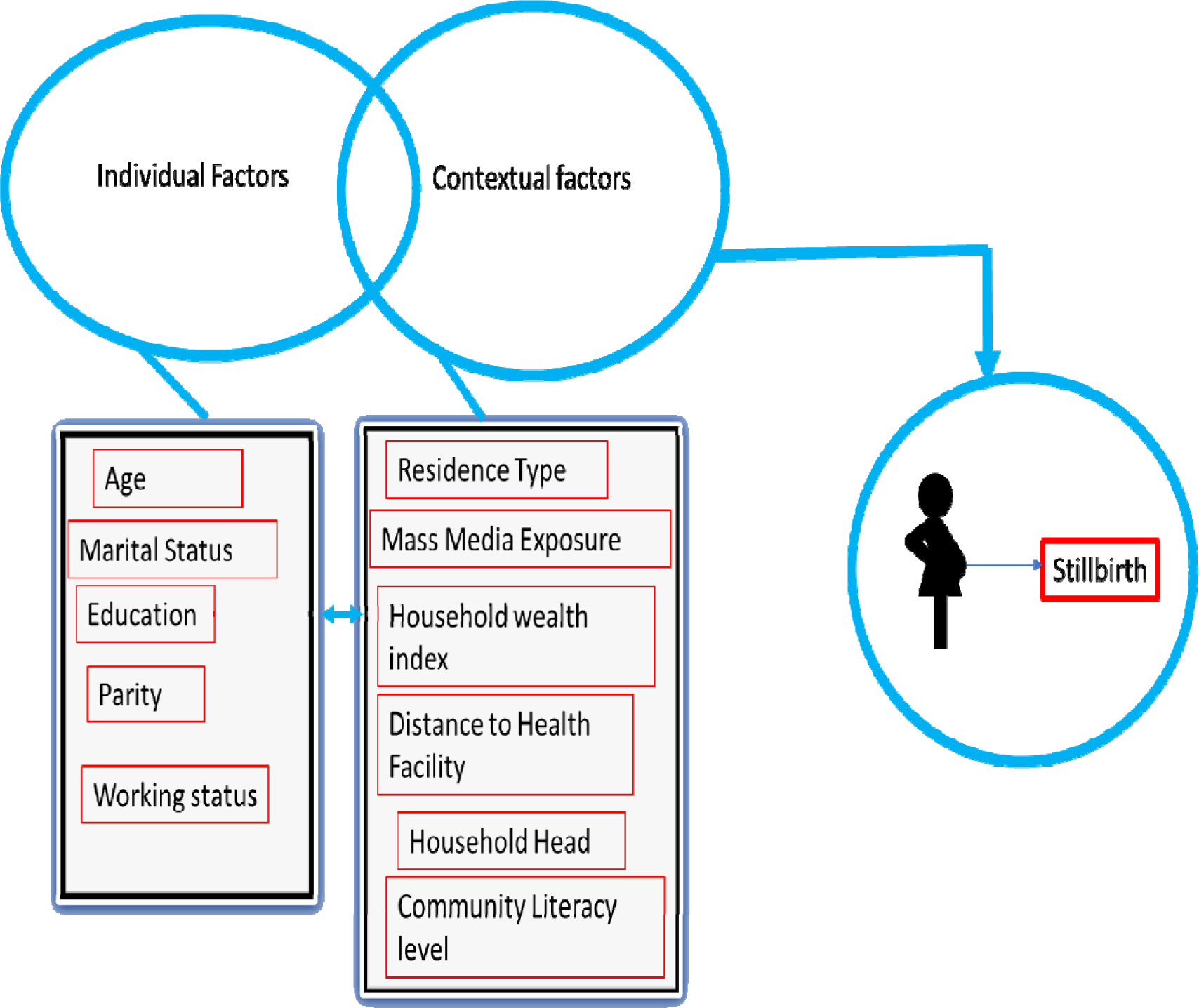
Factors influencing stillbirth at individual and contextual levels of the socioecological model.

**Table 1.** Description of study countries, samples, and region (N = 197, 329)

| **Selected DHS Countries, Survey Year, Sample Sizes, and Percentage** |  |  |  |  |
| --- | --- | --- | --- | --- |
| **Country** | **Region** | **Survey Year** | **Sample Size** | **Percentage** |
| Angola | Central Africa | 2015-2016 | 8936 | 4· 53 |
| Burkina Faso | West Africa | 2021 | 9284 | 4· 7 |
| Benin | West Africa | 2017-2018 | 8849 | 4· 48 |
| Burundi | East Africa | 2016-2017 | 8631 | 4· 37 |
| Ethiopia | East Africa | 2016 | 7193 | 3· 65 |
| Gabon | Central Africa | 2019-2021 | 2972 | 1· 51 |
| Ghana | West Africa | 2022-2023 | 6965 | 3· 53 |
| Gambia | West Africa | 2019-2020 | 5799 | 2· 94 |
| Guinea | West Africa | 2018 | 5504 | 2· 79 |
| Kenya | East Africa | 2022 | 7550 | 3· 83 |
| Comoros | East Africa | 2012 | 1965 | 1 |
| Liberia | West Africa | 2019-2020 | 4260 | 2· 16 |
| Lesotho | Southern Africa | 2014 | 2596 | 1· 32 |
| Madagascar | South-East Africa | 2021 | 9315 | 4· 72 |
| Mali | West Africa | 2018 | 6368 | 3· 23 |
| Mauritania | West Africa | 2019- 2021 | 7533 | 3· 82 |
| Malawi | East Africa | 2015- 2016 | 13448 | 6· 82 |
| Mozambique | East Africa | 2011 | 3581 | 1· 81 |
| Nigeria | West Africa | 2018 | 15632 | 7· 92 |
| Niger | West Africa | 2012 | 7617 | 3· 86 |
| Namibia | Southern Africa | 2013 | 3949 | 2 |
| Rwanda | East Africa | 2019- 2020 | 6167 | 3· 13 |
| Sierra Leone | West Africa | 2019 | 7374 | 3· 74 |
| Senegal | West Africa | 2015 | 4305 | 2· 18 |
| Tanzania | East Africa | 2022 | 7681 | 3· 89 |
| Uganda | East Africa | 2016 | 10255 | 5· 2 |
| South Africa | Southern Africa | 2016 | 1457 | 0· 74 |
| Zambia | Southern Africa | 2018 | 7369 | 3· 73 |
| Zimbabwe | Southern Africa | 2015 | 4774 | 2· 42 |
| Total |  |  | 197329 | 100 |

### Outcome variable

The primary outcome was stillbirth, defined as pregnancies lasting at least 28 weeks that ended in a fetal death^.1^ Birth outcome refers to whether a woman had a stillbirth or a live birth in her most recent pregnancy. We input all births within the prior five years by looping through the calendar, summing births, non-live pregnancies, and stillbirths from the births reported in the DHS. A loop iterates the entire (vcal_1) variable (calendar information string) within a five-year window. Within the loop, if the current position substring matches the code for stillbirth (“TPPPPPP”), the ‘stillbirths’ count was incremented by 1 (https://www.dhsprogram.com/data/calendar-tutorial/). We restricted our sample to women who gave birth within the last five years (v208), with stillbirth as the outcome variable. Stillbirth was dichotomized: 0 (“no stillbirth”) for absence of stillbirth, and 1 (“stillbirth”) for presence of stillbirth.

The observed stillbirth rate (SBR) was calculated as the ratio of reported stillbirths to total births (including both stillbirths and live births): SBR = sb/ (sb + lb) *1000 live births within a given period. Here, “sb” refers to the number of stillbirths ≥ 28 weeks of gestational age, and “lb” refers to the number of live births regardless of gestational age or birthweight.^19^

### Explanatory variables

We examined two groups of independent variables, considering the hierarchical structure of DHS data, where women are nested within clusters or communities. Following a comprehensive literature review, we selected individual — and contextual-level explanatory variables available in the DHS datasets. ^20,21^ We included eleven variables in the analysis and categorized them into individual- and contextual-level factors.

Individual factors considered were age, marital status, education, parity, and working status. We categorized age range as 15-19, 20-24, 25-29, 30-34, 35-39, 40-44, or 45-49. Education level was ‘no formal education,’ ‘primary,’ ‘secondary,’ or ‘higher.’ Another essential individual-level factor included in this study was parity, with four categories: number of children (1, 2, 3, or ≥4) ^22^; marital status (‘never married,’ ‘married,’ ‘cohabitation,’ ‘widowed,’ ‘divorced,’ or ‘separated’); current working status (‘no’ or ‘yes’). These categories were pre-coded in the DHS dataset.

Contextual factors included mass media exposure, including frequency of watching television, reading newspapers/magazines, and listening to the radio. Each mass media variable was categorized as ‘no exposure’ (not at all), ‘low exposure’ (less than once a week or at least once a week), or ‘high exposure’ (almost every day). ^23^ The household wealth index was based on the DHS measure of items available in each household. We used principal component analysis (PCA) ^24^ to group the available items into ‘poorest,” poorer,’ ‘middle,’ ‘richer,’ or ‘richest,’ representing the household wealth index.^23^ Place of residence (rural or urban) and travel to the health facility (‘big problem getting to the facility’ or ‘not a problem getting to the facility’) were pre-coded in the DHS dataset. Community-level female literacy was categorized into low, middle, or high literacy levels.^25^ Drawing on the Socioecological Model,^26^ **Figure 2** illustrates how complex interactions between individual and contextual factors at various levels affect stillbirth.

### Statistical analyses

We weighted the dataset per DHS guidelines (v005/1 000 000) to account for the complex sampling structure of the data and used the survey command (svy) in Stata.^17^ Sample weight equalization gave equal weight to each survey, so if one survey had a large sample, it did not predominate the pooled results. This process ensured that the sample weights aligned with each country’s clusters and strata. We used STATA software, version 18·0 SE (Stata Corporation, TX, USA) for weighted data analysis.^27^ We appended all data from SSA DHS after extracting the essential variables. In our analyses, we included participants with complete information on all covariates but excluded a “missing” category.

We sampled sociodemographic variables to generate represented percentages for each country. The weighted datasets for the 29 countries were merged to develop the overall prevalence of stillbirth among women across the study. We used a map to visually represent the varying prevalence of stillbirth by country (**Figure 3**). We used bivariate analysis to examine the relationship between stillbirth and the explanatory variables. The Pearson chi-square test of independence provided information on the distribution of stillbirth rates across the different variables and indicated any significant associations. We checked for multicollinearity among the explanatory variables using the variance inflation factor (VIF). The results showed no evidence of high collinearity (maximum VIF is 2·07, the minimum VIF is 1·03, mean VIF is 1· 38, and condition number is 30·02).

**Figure 3.**
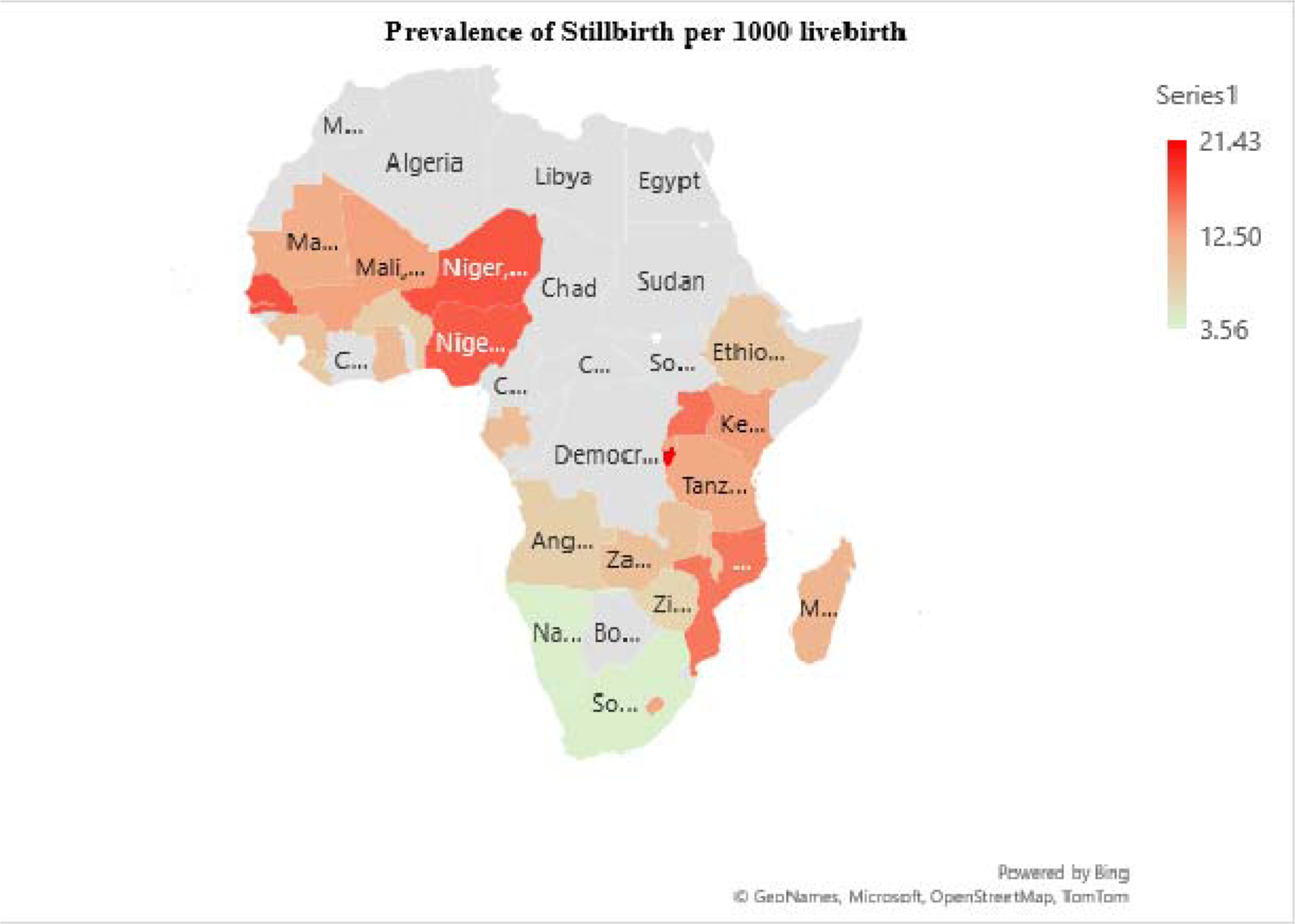
Prevalence of stillbirth in Sub-Saharan African countries.

Finally, we analyzed multilevel binary logistic regression using four models (Model O-III). Model O was an empty model without any explanatory variables, serving as a baseline. Model I included only individual-level variables, Model II had only contextual variables, and Model III, the complete model, incorporated individual and contextual-level variables. The regression analysis results were presented as adjusted odds ratios (aOR) with corresponding 95% confidence intervals (CI), and statistical significance was set at p < 0·001 and p < 0·005, indicating associations between the explanatory variables and stillbirth. The analysis accounted for non-response and under-sampling by applying the survey sample weight. We used the Stata command ‘melogit’ to fit these models, as well as the log-likelihood ratio and Akaike’s information criterion (AIC) tests for model comparison.

### Ethical considerations

This study was based on a secondary dataset with no identified participant information. The authors obtained and were approved to use the dataset by MEASURE DHS.

## Findings

### Prevalence of stillbirth in SSA

The sample was 197,329 DHS participants from 29 SSA countries (**Table 1**). The prevalence of stillbirth among women who had given birth within the last five years preceding the interview in SSA was 319·06 per 1000 live births. Burundi recorded the highest prevalence, 21·4/1000 live births, whereas Comoros recorded the lowest, 3·6/1000 live births (**Figure 3**).

### Individual-level predictors of stillbirth among women in sub-Saharan Africa

Age, education, parity, and marital status were significantly associated with a stillbirth at p<0· 001 (**Table 2**). Several factors were associated with an increased risk of stillbirth among women in SSA countries (**Table 3**). There was no significant difference in the odds of stillbirth among women in age categories 15-19 and 20-24 compared to those in the 45-49 age category. However, the odds of stillbirth were higher in those aged 25-29, 30-34, 35-39, and 40-44 (aOR=1·98, 95%CI: 1·26 to 3·11), (aOR=2·14, 95%CI: 1·37 to 3·33), (aOR=2·29, 95%CI: 1·45 to 3·60), and (aOR=1·99, 95%CI: 1·24 to 3·50) respectively (**Figure 4**). Maternal education was negatively associated with the risk of stillbirth. Compared to women with no education, higher maternal education levels were associated significantly with decreased risk of stillbirth with coefficients ranging from 64% to 90% (aOR=0· 90 95% CI: 0·80 to 1·00, and aOR=0·64 95% CI: 0·45 to 0·90 respectively) (**Table 3**). While single (never in a union) women had significantly lower odds of stillbirth (aOR=0·38, 95% CI: 0·25 to 0·58), married or cohabitating women had higher odds of stillbirth (aOR=1·40, 95% CI: 1·04 to 1·88) and (aOR=1·39, 95% CI: 1· 02 to 1· 90) respectively. Women with higher parity had significantly higher odds of experiencing stillbirth in women with two births (aOR= 0·38,95% % CI of 0·32 to 0·46) or three births (aOR= 0·31,95%CI of 0·26 to 0· 59), compared to women with one birth (**Table 3**). Women with a job had increased risk (aOR 1· 20 95%CI:1· 08 to 1· 35) compared to women without a job.

**Figure 4.**
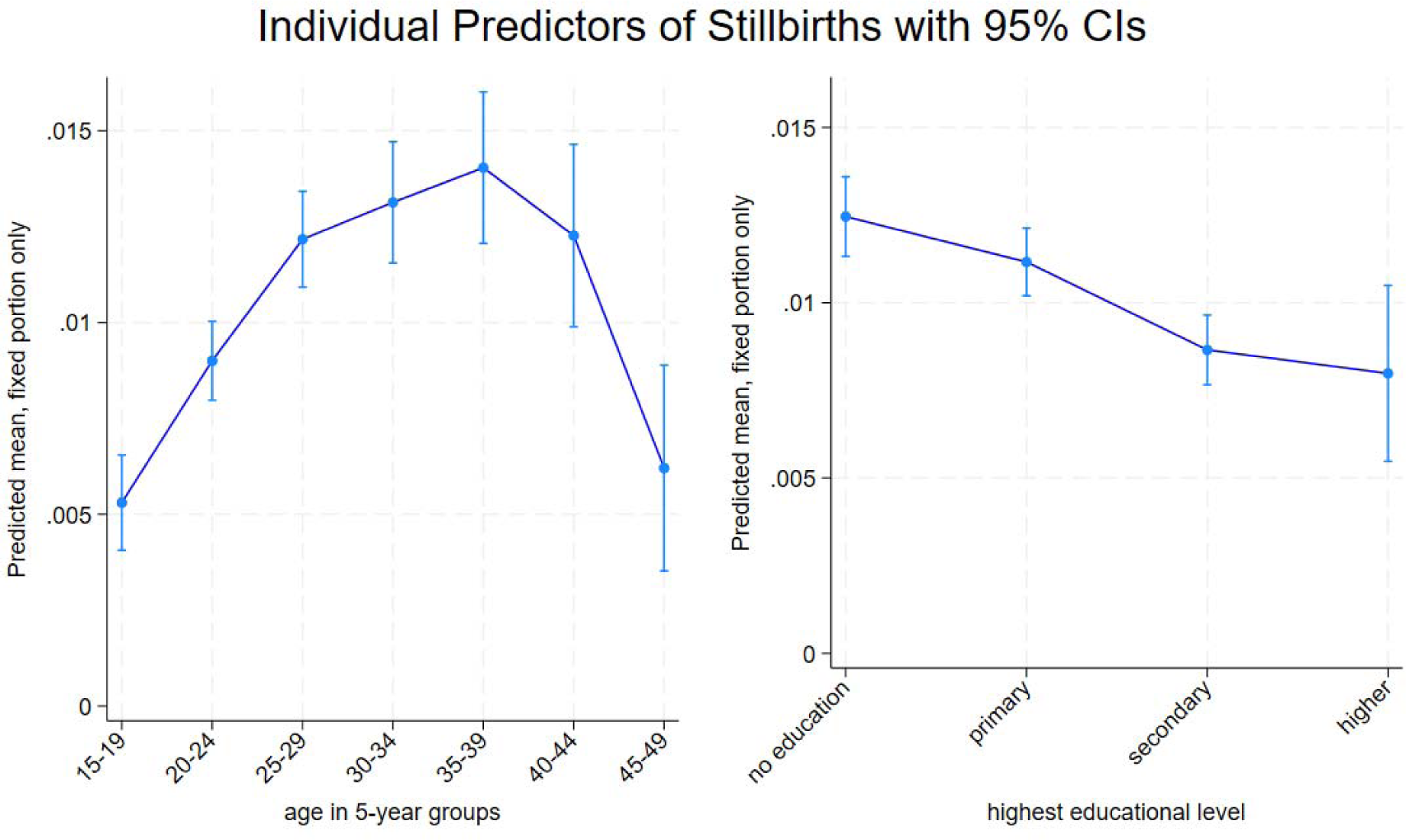
Individual-level predictors of stillbirth.

**Table 2.**
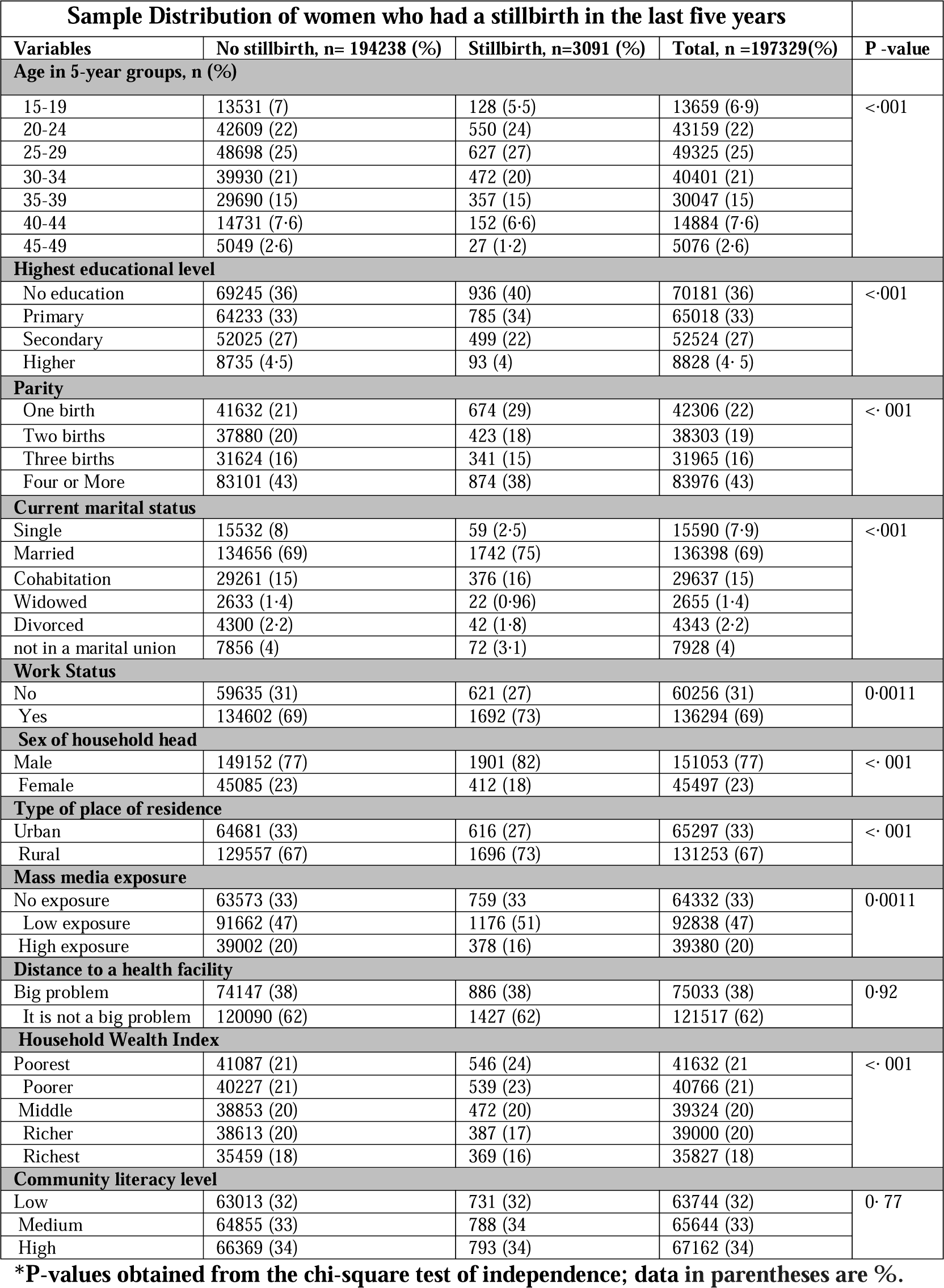
Association between stillbirth and explanatory variables.

| **Sample Distribution of women who had a stillbirth in the last five years** |  |  |  |  |
| --- | --- | --- | --- | --- |
| **Variables** | **No stillbirth, n= 194238 (%)** | **Stillbirth, n=3091 (%)** | **Total, n =197329(%)** | **P -value** |
| **Age in 5-year groups, n (%)** |  |  |  |  |
| 15-19 | 13531 (7) | 128 (5.5) | 13659 (6.9) | < 001 |
| 20-24 | 42609 (22) | 550 (24) | 43159 (22) |  |
| 25-29 | 48698 (25) | 627 (27) | 49325 (25) |  |
| 30-34 | 39930 (21) | 472 (20) | 40401 (21) |  |
| 35-39 | 29690 (15) | 357 (15) | 30047 (15) |  |
| 40-44 | 14731 (7.6) | 152 (6.6) | 14884 (7.6) |  |
| 45-49 | 5049 (2.6) | 27 (1.2) | 5076 (2.6) |  |
| **Highest educational level** |  |  |  |  |
| No education | 69245 (36) | 936 (40) | 70181 (36) | < 001 |
| Primary | 64233 (33) | 785 (34) | 65018 (33) |  |
| Secondary | 52025 (27) | 499 (22) | 52524 (27) |  |
| Higher | 8735 (4.5) | 93 (4) | 8828 (4.5) |  |
| **Parity** |  |  |  |  |
| One birth | 41632 (21) | 674 (29) | 42306 (22) | < 001 |
| Two births | 37880 (20) | 423 (18) | 38303 (19) |  |
| Three births | 31624 (16) | 341 (15) | 31965 (16) |  |
| Four or More | 83101 (43) | 874 (38) | 83976 (43) |  |
| **Current marital status** |  |  |  |  |
| Single | 15532 (8) | 59 (2.5) | 15590 (7.9) | < 001 |
| Married | 134656 (69) | 1742 (75) | 136398 (69) |  |
| Cohabitation | 29261 (15) | 376 (16) | 29637 (15) |  |
| Widowed | 2633 (1.4) | 22 (0.96) | 2655 (1.4) |  |
| Divorced | 4300 (2.2) | 42 (1.8) | 4343 (2.2) |  |
| not in a marital union | 7856 (4) | 72 (3.1) | 7928 (4) |  |
| **Work Status** |  |  |  |  |
| No | 59635 (31) | 621 (27) | 60256 (31) | 0.0011 |
| Yes | 134602 (69) | 1692 (73) | 136294 (69) |  |
| **Sex of household head** |  |  |  |  |
| Male | 149152 (77) | 1901 (82) | 151053 (77) | < 001 |
| Female | 45085 (23) | 412 (18) | 45497 (23) |  |
| **Type of place of residence** |  |  |  |  |
| Urban | 64681 (33) | 616 (27) | 65297 (33) | < 001 |
| Rural | 129557 (67) | 1696 (73) | 131253 (67) |  |
| **Mass media exposure** |  |  |  |  |
| No exposure | 63573 (33) | 759 (33) | 64332 (33) | 0.0011 |
| Low exposure | 91662 (47) | 1176 (51) | 92838 (47) |  |
| High exposure | 39002 (20) | 378 (16) | 39380 (20) |  |
| **Distance to a health facility** |  |  |  |  |
| Big problem | 74147 (38) | 886 (38) | 75033 (38) | 0.92 |
| It is not a big problem | 120090 (62) | 1427 (62) | 121517 (62) |  |
| **Household Wealth Index** |  |  |  |  |
| Poorest | 41087 (21) | 546 (24) | 41632 (21) | < 001 |
| Poorer | 40227 (21) | 539 (23) | 40766 (21) |  |
| Middle | 38853 (20) | 472 (20) | 39324 (20) |  |
| Richer | 38613 (20) | 387 (17) | 39000 (20) |  |
| Richest | 35459 (18) | 369 (16) | 35827 (18) |  |
| **Community literacy level** |  |  |  |  |
| Low | 63013 (32) | 731 (32) | 63744 (32) | 0.77 |
| Medium | 64855 (33) | 788 (34) | 65644 (33) |  |
| High | 66369 (34) | 793 (34) | 67162 (34) |  |

**\*P-values obtained from the chi-square test of independence; data in parentheses are %.**

**Table 3.** Factors associated with stillbirth among women in sub-Saharan Africa.

| Variables | Model O<br>Empty model | Model I<br>aOR [95% CI] | Model II<br>aOR [95% CI] | Model III<br>aOR [95% CI] |
| --- | --- | --- | --- | --- |
| **Age** |  |  |  |  |
| 15-19 |  | 0.93 [0.56,1.54] |  | 0.85[0.52,1.41] |
| 20-24 |  | 1.55 [0. 98,2. 43] |  | 1.46 [0.93,2.29] |
| 25-29 |  | 2.03**[1. 29,3.19] |  | 1.98** [1.26,3.11] |
| 30-34 |  | 2.14***[1. 37,3.33] |  | 2. 14***[1. 37,3. 33] |
| 35-39 |  | 2.26***[1.43,3.56] |  | 2. 29***[1. 45,3. 60] |
| 40-44 |  | 1. 98** [1.23,3. 17] |  | 1. 99** [1.24,3. 20] |
| 45-49[ref] |  | 1[1. 00,1. 00] |  | 1[1. 00,1. 00] |
| **Education** |  |  |  |  |
| No Education [ref] |  | 1[1. 00,1. 00] |  | 1[1. 00,1. 00] |
| Primary |  | 0. 88*[0. 78,0. 98] |  | 0. 90[0. 80,1. 00] |
| Secondary |  | 0. 61***[0. 53,0. 71] |  | 0. 69***[0. 59,0. 80] |
| Higher |  | 0. 52***[0. 38,0. 71] |  | 0. 64** [0. 45,0. 90] |
| **Marital Status** |  |  |  |  |
| Single |  | 0. 38***[0. 25,0. 58] |  | 0. 38***[0. 25,0. 58] |
| Married |  | 1. 51** [1. 14,2. 01] |  | 1. 40* [1. 04,1. 88] |
| Cohabitation |  | 1. 48* [1. 09,2. 00] |  | 1. 39*[1. 02,1. 90] |
| Widowed |  | 1. 07 [0. 59,1. 96] |  | 1. 09[0. 60,2. 00] |
| Divorced |  | 1. 05 [0. 68,1. 62] |  | 1. 05[0. 68,1. 62] |
| Not in marital union[ref] |  | 1[1. 00,1. 00] |  | 1[1. 00,1. 00] |
| **Parity** |  |  |  |  |
| One[ref] |  | 1[1. 00,1. 00] |  | 1[1. 00,1. 00] |
| Two |  | 0. 48***[0. 41,0. 57] |  | 0. 48***[0. 41,0. 56] |
| Three |  | 0. 39***[0. 33,0. 47] |  | 0. 38***[0. 32,0. 46] |
| Four+ |  | 0. 33***[0. 27,0. 39] |  | 0. 31 ***[0. 26,0. 37] |
| **Work Status** |  |  |  |  |
| No[ref] |  | 1[1. 00,1. 00] |  | 1[1. 00,1. 00] |
| Yes |  | 1. 23***[1. 10,1. 38] |  | 1. 20** [1. 08,1. 35] |
| **Mass Media Exposure** |  |  |  |  |
| No exposure |  |  | 1. 045 [0. 88,1. 24] | 1. 01 [0. 85,1. 22] |
| Low Exposure |  |  | 1. 184* [1. 02,1. 38] | 1. 16 [0. 99,1. 35] |
| High Exposure[ref] |  |  | 1[1. 00,1. 00] | 1[1. 00,1. 00] |
| **Residence type** |  |  |  |  |
| Urban [ref] |  |  | 1[1. 00,1. 00] | 1[1. 00,1. 00] |
| Rural |  |  | 1. 29***[1. 12,1. 48] | 1. 22** [1. 06,1. 41] |
| **Wealth index** |  |  |  |  |
| Poorest |  |  | 1. 11 [0. 90,1. 35] | 1. 19 [0. 96,1. 46] |
| Poorer |  |  | 1. 10 [0. 91,1. 34] | 1. 18 [0. 96,1. 44] |
| Middle |  |  | 1. 01 [0. 83,1. 23] | 1. 09 [0. 88,1. 33] |
| Richer |  |  | 0. 89 [0. 74,1. 08] | 0. 93[0. 77,1. 13] |
| Richest[ref] |  |  | 1 [1. 00,1. 00] | 1 [1. 00,1. 00] |
| **Distance to facility** |  |  |  |  |
| Big Problem [ref] |  |  | 1[1.00,1. 00] | 1[1.00,1. 00] |
| Not a problem |  |  | 1. 07 [0.96,1. 19] | 1. 05 [0.94,1. 17] |
| **Household head** |  |  |  |  |
| Male [ref] |  |  | 1[1.00,1.00] | 1[1.00,1.00] |
| Female |  |  | 0.74***[0.65,0. 83] | 0. 86* [0. 75,0.99] |
| **Community literacy level** |  |  |  |  |
| Community literacy level |  |  | 0. 92 [0. 81,1.05] | 0. 92 [0. 80,1. 05] |
| Community literacy level |  |  | 0. 99 [0. 87,1.12] | 0. 98 [0. 86,1. 12] |
| Community literacy level[ref] |  |  | 1 [1. 00,1. 00] | 1[1. 00,1. 00] |
| **Random Effect Model** |  |  |  |  |
| PSU 95% variance | 1. 21***[1. 14,1. 28] | 1. 21***[1. 14,1. 29] | 1. 21***[1. 14,1. 28] | 1. 21***[1. 14,1. 28] |
| ICC | 0.055 | 0.055 | 0.054 | 0. 054 |
| Walden chi-square | Reference | 343. 68 | 63. 9 | 398. 38 |
| **Model fitness** |  |  |  |  |
| Log-likelihood | -12526. 37 | -12294. 81 | -12473. 58 | -12265. 566 |
| BIC | 25077. 13 | 24833. 46 | 25105. 66 | 24909. 1 |
| AIC | 25056. 74 | 24629. 61 | 24973. 15 | 24593. 13 |
| *N* | 197329 | 197329 | 197329 | 197329 |
| Number of clusters | 1,683 | 1,683 | 1,683 | 1,683 |
| Exponentiated coefficients; 95% confidence intervals in brackets; *$p < 0.05$, **$p < 0.01$, ***$p < 0.001$<br>PSU Primary Sampling Unit,<br>ICC Intraclass correlation coefficient, LR Test Likelihood ratio Test, AIC Akaike's Information Criterion, aOR adjusted Odds Ratios, CI Confidence Interval<br>Model 0 is the Empty model, a baseline model without any explanatory variable.<br>Model 1 is adjusted for the individual level.<br>Model 2 is adjusted for contextual-level variables.<br>Model 3 is the final model adjusted for all explanatory variables and survey country. | | | | |

### Contextual (household and community-level) predictors of stillbirth

Place of residence, sex of household head, and household wealth index were significantly associated with a stillbirth at p<0· 001 (**Table 2**). Living in a rural residential area was a significant risk factor (aOR=1·22 95%CI:1·06 to1·41). Exposure to mass media did not significantly affect stillbirth (aOR 1·01, 95% CI 0· 85to 1·22). When the household head was female, the odds of stillbirth were significantly lower (aOR=0·86, 95% CI: 0·75 to 0·99) compared to households headed by males. Community literacy level was not a significant predictor of stillbirth (**Table 3**).

### Random effects (measures of variation) results

The random effects model (**Table 3**) indicated a statistically significant variation in determinants of stillbirth across the clusters. In the empty model, there were substantial variations in the likelihood of determinant of stillbirth across the clustering of the primary sampling units (PSUs;1·21(95% CI 1·14 to 1·28). The empty model’s intraclass correlation (ICC) value showed 0·055 of the variation in stillbirth rates attributed to the between-cluster variations of the characteristics. The between-cluster difference dropped to 0·055 in Model I, to 0·054 in Model II, and maintained its value 0·055 in Model III.

These ICC results suggested that the variations in the likelihood of experiencing stillbirth could be attributed to the variances across the clusters. The AIC values exhibited a similar U-shaped pattern as the ICC values, reaching their lowest point in Model III. Therefore, we chose Model III as the most suitable model to analyze the determinants of stillbirth (AIC; 24593·13).

## Interpretation

The study provides insights into the multifaceted nature of stillbirth risk factors in SSA and highlights the influence of individual factors on prevalence. Maternal age, work status, rural residence, and education were significant determinants, with older age and less education associated with higher risk. Mass media exposure did not significantly impact stillbirth odds or community literacy.

At the individual level, stillbirth odds increased with age and parity. This finding was consistent with a study in the United States showing a four-fold-increased risk of stillbirth in women ≥40 years compared to women aged 20-29 years.^28–30^ Associated physiological changes in the reproductive system, underlying medical conditions, or decreased fertility with advancing age could explain this.^29^ Previous studies found an association between age and parity. ^31,32^ In an SSA context, stillbirth might be associated with increasing parity with increased age if age and parity were correlated. It is crucial to support pregnant older women with comprehensive management strategies, including preconception counseling, regular monitoring, collaboration with specialized providers, risk assessments, fetal health monitoring, and education on warning signs. This holistic approach addresses the unique challenges associated with pregnancies and could reduce the risk of stillbirth.

Additionally, our study found that higher maternal education was associated with a lower risk of stillbirth among women, which is consistent with the results of other studies in different regions and SSA.^5^ A study in Northern Tanzania, East Africa, and India found that stillbirth risk was significantly associated with lower maternal education.^7,31,33^ This pattern can be attributed to the influence of education on socioeconomic status, access to healthcare, and maternal health knowledge in identifying the signs of stillbirth. Some strategies to increase health literacy are offering information in preferred languages, utilizing technology tailored to lower literacy levels, and engaging community Health Workers (CHWs).^34,35^

Rural residents had a higher risk of stillbirth than women who lived in urban areas. This is consistent with research in India, which identified rural residence as a strong risk factor for stillbirth, with women in rural areas having 27% higher odds of stillbirth compared to women in urban areas.^31^ The impact of rural residence on stillbirth risk underscores the disparities in healthcare access, infrastructure, and socioeconomic conditions between urban and rural settings, potentially influencing pregnancy outcomes.

Further, while the reasons remain unclear, the female head of household had a slightly lower risk of stillbirth than those with a male head. This finding is consistent with a study that identified a female head of household as a protective factor for stillbirths, potentially influencing the associations between risk factors and stillbirth outcomes.^36^ Increased autonomy in healthcare choices, access to resources and information, and more robust social support networks could all contribute to this observed difference. Further research is needed, but understanding the link between gender roles and pregnancy outcomes could inform policies and interventions to support all women and families better.

### Implications for practice

The study’s findings highlight the importance of interventions to reduce stillbirth rates, particularly among older women, women with lower levels of education, and women living in rural areas. Interventions could include providing education about the signs of stillbirth, improving access to quality healthcare, and promoting safe birth practices. At the community level, awareness campaigns and educational programs can empower women with knowledge about prenatal care and maternal health. In addition, the study’s findings support broader national and international strategies to improve maternal and child health to achieve Sustainable Development Goal 3, which focuses on ensuring healthy lives and promoting well-being for all ages.

### Strengths and limitations

The major strengths of this study were the use of recent nationally representative DHS data from 29 SSA countries and the adoption of a multilevel approach to the analyses. This approach allowed for examining associations between stillbirth and individual and contextual-level factors, providing a more comprehensive understanding of the complex determinants of stillbirth across different geographic contexts. Notably, this study serves as the first comprehensive analysis of stillbirth risk factors pooling data from such a large and diverse group of SSA countries.

We acknowledge some limitations. The data was cross-sectional; therefore, no causality could be inferred. While the DHS dataset is large and nationally representative, the stillbirth sample size was relatively small, particularly in specific SSA countries. This limited the statistical power to detect associations between potential determinants and stillbirth. Further research and longitudinal studies are needed to establish a more definitive understanding of the determinants of stillbirth in this context.

### Conclusion

Our results indicate that individual characteristics and some social and environmental factors contribute to stillbirth risk. Our findings can inform the development of interventions to reduce stillbirth rates in this region. Policy, community, and health system modifications are imperative to enhance birth outcomes among women in SSA. Health systems should prioritize the development of comprehensive, culturally sensitive maternal health programs that address the unique challenges faced by women in SSA, thereby contributing to improved birth outcomes and reducing stillbirths.

## Abbreviations

SSA: Sub-Saharan Africa
SDG: Sustainable Development Goal
DHS: Demographic and Health Surveys
aOR: Adjusted odds ratio.

## Acknowledgments

The authors are grateful to MEASURE DHS for granting access to the dataset used in this study.

## Author contributions

KKA, OO, and FI contributed to conceptualizing the study. KKA, OO, YYK, AG, and FI reviewed the literature. KKA performed the analysis and reviewed it by OO. OO, AG, and FI supervised and contributed intellectually through the development of the manuscript. They provided technical support and critically reviewed the manuscript for its intellectual content. All authors read and amended drafts of the paper and approved the final version.

## Data availability

The datasets utilized in this study can be accessed from the DHS Program and are available at https://dhsprogram.com/data/available-datasets.cfm

